# Identifying Plasma Proteins Associated with Risk of Solid Cancers: A 25-Year Prospective Analysis of 4,712 Circulating Proteins in the ARIC Study

**DOI:** 10.64898/2026.03.16.26348527

**Authors:** Ziqiao Wang, Vernon A. Burk, Zhe Huang, Hana Zahed, David C. Muller, James Yarmolinsky, Matthew A. Lee, Corinne E. Joshu, Zhike (Coco) Lin, Anna Prizment, Kenneth R. Butler, David J. Couper, Karl Smith-Byrne, P. Martijn Kolijn, Roel C.H. Vermeulen, Elio Riboli, Marc J. Gunter, Josef Coresh, Nilanjan Chatterjee, Elizabeth A. Platz

## Abstract

This study investigated potential pre-diagnostic proteomic risk markers for 7 solid cancers independent of known risk factors within a multi-center, prospective cohort study. Using the SomaScan® 5K assay, we analyzed 4,712 unique plasma proteins (4,955 aptamers) in the Atherosclerosis Risk in Communities (ARIC) study among 9,391 middle-aged and older Black (23%) and White (77%) men and women. Over a maximum follow-up of 25.9 years, incident cases included 136 bladder, 271 colorectal, 96 kidney, 22 liver, 416 lung, 88 pancreatic, and 588 prostate cancers. After false discovery rate (FDR) correction, we identified 144 unique protein-cancer associations in common risk-factor adjusted models, and 41 protein-cancer associations in both common and cancer site-specific risk-factor adjusted models. Associations included several novel circulating proteins related to liver (33 proteins) and lung (4 proteins) cancer risk, and confirmed previously established proteins associated with kidney (HAVCR1 and MMP7) and prostate (KLK3 and ACP3) cancer risk. External validation in the European Prospective Investigation into Cancer and Nutrition cohort (SomaScan 7K) confirmed that the majority of FDR-significant proteins showed consistent effect directions and nominal significance, with the proportion of confirmed proteins varying between 75% and 100% depending on the cancer site. Time-lagged analysis demonstrated that 90% of the identified cancer-associated proteins are markers for long-term cancer risk, with observed associations more than 5 years pre-diagnosis after multiple-testing correction. These findings underscore the potential of circulating proteomic markers beyond known risk factors for elucidating etiologic mechanisms and improving risk stratification across cancers.

## Introduction

Cancer is the second leading cause of death in the United States, with approximately one in three individuals expected to be diagnosed with the disease at some point in their lifetime. In 2026, it is estimated that more than 2 million new cancer cases will be diagnosed nationwide.^1^ Public health strategies to reduce the cancer burden in populations include identifying biomarkers for early detection, risk factors that can help to define preventive interventions, and at-risk populations who will benefit the most from certain interventions. Epidemiologic studies over many decades have investigated the association of lifestyle and environmental factors, as well as family and medical history, with the risk of different cancers. Large-scale genomic studies conducted over the past two decades have identified hundreds of cancer susceptibility loci across different cancer sites, leading to the development of polygenic risk scores for cancer risk stratification.^2–10^ Recent advances in proteomic technologies now offer complementary opportunities to characterize proteins—which capture the combined influence of genetics and environment—as cancer risk factors and early detection biomarkers.

Two leading platforms that are currently implementing population-scale studies are Olink and SomaScan. Different platforms target different sets of proteins, but even for common proteins there are known differences in measurements across the 2 platforms due to differences in the technologies.^11–13^ Therefore, studies based on alternative platforms are needed for robust identification of proteomic risk factors and biomarkers for cancer risk. Multiple studies^14–16^ have now reported associations between a variety of proteins and cancer risk based on the large UK Biobank study, which has ascertained plasma levels of nearly 3,000 proteins based on the antibody-based Olink platform. We recently used data from the SomaScan platform available for the Atherosclerosis Risk in Communities (ARIC) study to identify that the plasma level of protein LEG1 homolog and adenosine diphosphate (ADP)-dependent glucokinase are associated with risk of post-menopausal breast cancer^17^ and confirmed these associations in the European Prospective Investigation into Cancer and Nutrition (EPIC), which also implemented the SomaScan platform. Similarly, a study in the EPIC cohort discovered hundreds of proteins associated with lymphoid cancers, and the top hits were largely confirmed in the ARIC and the UK Biobank studies.^18^ Robust identification of proteomic-cancer associations with broad public health utility requires studies involving individuals of diverse ancestry. However, large studies such as the UK Biobank and EPIC, have predominantly included individuals of European ancestry. The goal of our current study is to provide a comprehensive characterization of the association of SomaScan-measured plasma proteins with the risk of solid cancers among White and Black participants in the ARIC study, and to conduct confirmation analyses in the EPIC study, an independent cohort.

We conducted high-throughput proteomic profiling of 4,712 proteins (4,955 aptamers) quantified using the SomaScan® 5K assay and their associations with the future risk of 7 solid cancers, independent of known risk factors (**Fig. 1**). This analysis included a maximum follow-up of 25.9 years involving 9,391 White and Black male and female participants from the ARIC study. Proteins that remained significant after correction for multiple testing were subsequently evaluated in the EPIC study, which included comparable aptamer-based proteomic data and model adjustments. We further leveraged the long follow-up of the ARIC study to investigate how the strength of protein-cancer association varies over time since measurement, and longitudinal changes in protein levels across 3 study timepoints among cancer-free individuals over a 21-year observation period to understand the within-person stability of protein measurements over time. Finally, pathway enrichment analyses were performed to investigate potential biological mechanisms underlying proteins associated with liver cancer risk.

**Figure 1.**
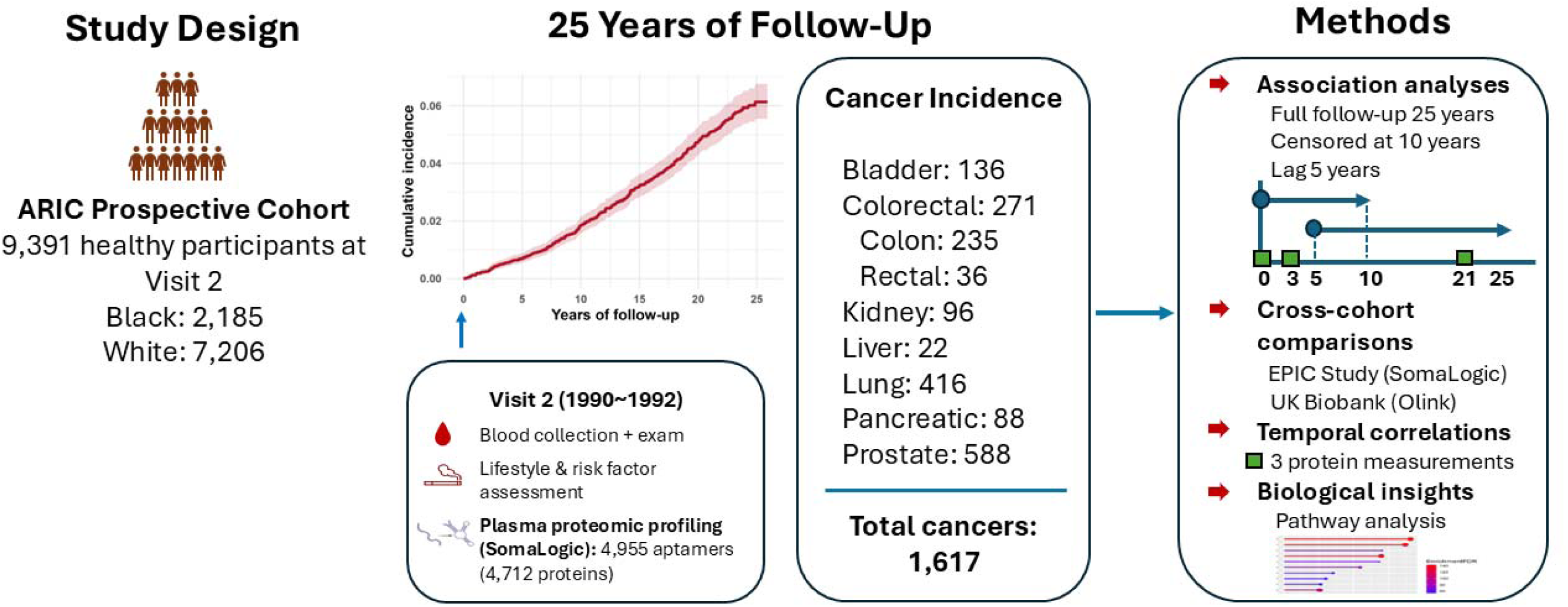
Graphical abstract illustrating the study design, data, and statistical methods for identifying plasma proteins associated with risk of solid cancers in the ARIC study.

**Figure 2.**
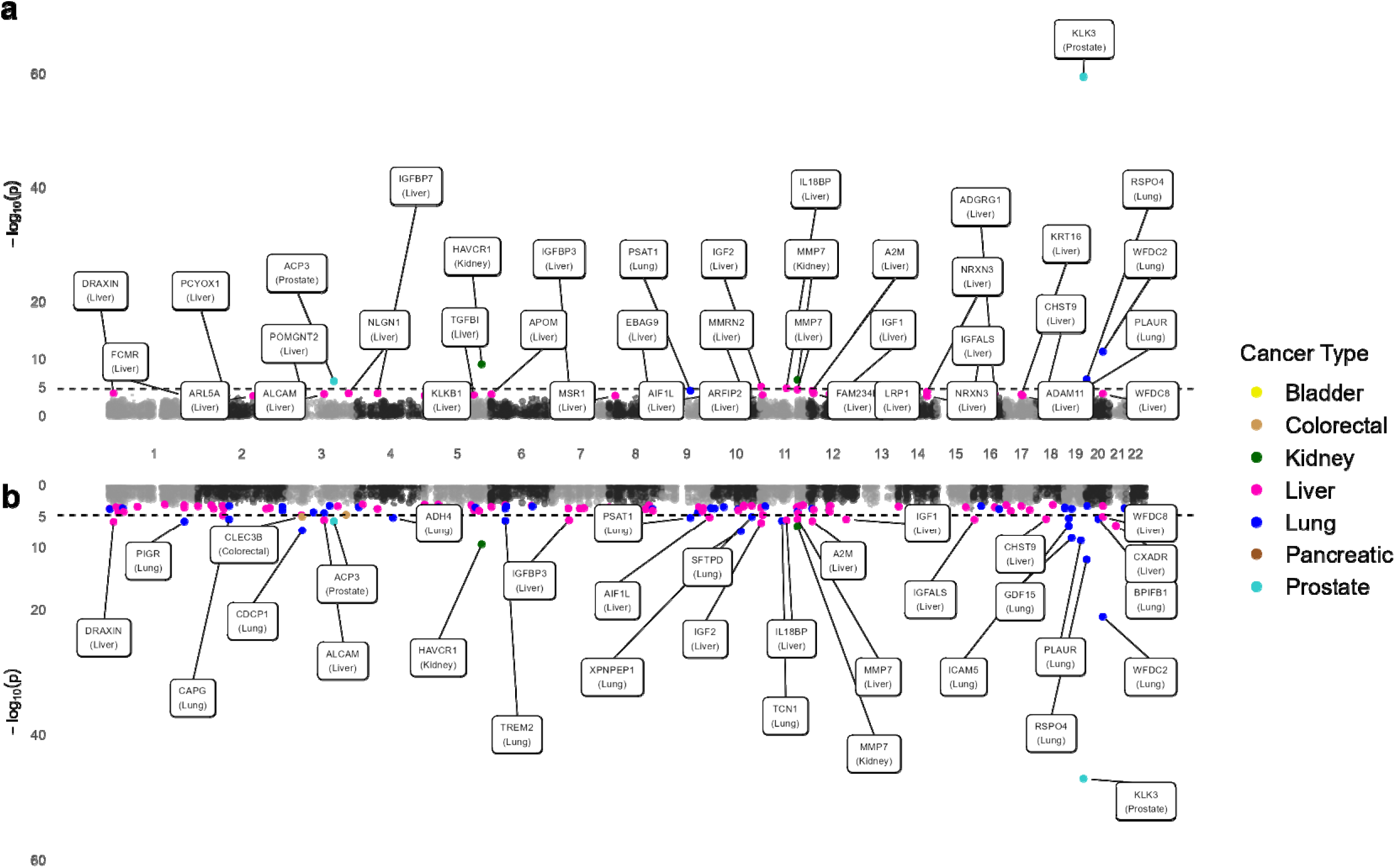
Miami plot of significant proteins among 4,955 aptamers for 7 cancers. (a) Adjusted for common and site-specific risk factors (Model 2; summary statistics in Table 1 and S7; covariates in Supplementary Table S2). (b) Adjusted for common cancer risk factors (Model 1; summary statistics in Table S6).

## Methods

### Study population

This study is based in the ARIC (RRID:SCR_021769) community-based, prospective cohort of 15,792 participants recruited from Forsyth County, North Carolina; Jackson, Mississippi; suburbs of Minneapolis, Minnesota; and Washington County, Maryland in the US.^19^ Of them, 55% were women and 73% were White. Participants were recruited for Visit 1 between 1987-1989. We included in the analytic cohort 11,798 participants who had plasma protein measured using blood collected in 1990-1992 (Visit 2), the earliest visit with proteomics data. We then excluded participants who did not provide consent for non-cardiovascular disease research or use of genetic information, who are not self-identified as Black or White, or who had any prevalent cancer history before Visit 1, or the date of censor or cancer diagnosis occurred before Visit 2, or who had missing covariate data (Visit 1, Visit 2). Following these exclusions, the analysis cohort included 9,391 participants (**Table S1**).

### Ascertainment of incident solid cancer cases

This study included 7 solid cancers: bladder, colorectal, kidney, liver, lung, pancreatic, and prostate. Primary liver cancer was included because the liver is the major producer of proteins in plasma, while the other 6 cancers were chosen due to having the highest incidence rates in the cohort compared to other cancers. Female breast cancer is not included in this work, as results for this site were recently reported.^17^ Incident primary cancer cases were ascertained between 1987 and 2015 through linkage to cancer registries in Maryland, Minnesota, Mississippi, and North Carolina.^20^ Cases were also ascertained following a participant report of a new cancer diagnosis on an annual or semi-annual (since 2012) call and confirmed by medical record review. Other cases were identified by abstraction of routinely collected hospital discharge summaries and medical records, and death certificates (cancer as the underlying cause). The cancers included in this analysis were identified using the following ICD codes: bladder cancer (ICD-O-1: 188.0-188.9; ICD-O-2: 67.0-67.9; ICD-O-3: C67.0-C67.9; ICD-8 & 9: 188.0-188.9; ICD-10: 67.0-67.9), colorectal cancer (ICD-O-1: 153.0-153.4, 153.6-153.8, 154.0-154.1; ICD-O-2: 18.0, 18.2-18.9, 19.9, 20.9; ICD-O-3: C18.0, C18.2-C18.9, C19.9, C20.9; ICD-8 & 9: 153.0-153.4, 153.6-153.8, 154.0-154.1; ICD-10: 18.0, 18.2-18.9, 19.9, 20.9), kidney cancer (ICD-O-1: 189; ICD-O-2: 64-66, 68; ICD-O-3: C64-66, 68; ICD-8 & 9: 189; ICD-10: 64-66, 68), liver cancer (ICDO-1: 155.0; ICD-O-2: 22.0; ICD-O-3: C22.0; ICD-8 & 9: 155.0; ICD-10: 22.0, 22.9), lung cancer (ICD-O-1: 162.0-162.9; ICD-O-2: 34.0-34.9; ICD-O-3: C34.0-C34.9; ICD-8 & 9: 162.0-162.9; ICD-10: 34.0-34.9), pancreatic cancer (ICD-O-1: 157.0–9; ICD-O-2: 25.0–9; ICD-O-3: C25.0–9; ICD-8: 157.0–9; ICD-9: 157.0–9; ICD-10: 25.0–9), prostate cancer (ICD-O-1: 185; ICD-O-2: 61.9; ICD-O-3: C61.9; ICD-8: 185; ICD-9: 185; ICD-10: 61.9). Follow-up person-years were calculated from Visit 2 until the first diagnosis of primary incident cancer, death, or censoring date (December 31, 2015), whichever came first.

### Plasma protein measurement, preprocessing, and normalization

At Visit 2, blood samples were collected, and plasma was aliquoted using a standardized protocol, frozen at -80°C, and transferred to the ARIC central laboratory. For the proteomics assay, plasma was thawed, aliquoted into barcoded microtiter plates, and shipped to SomaLogic, where the relative fluorescence unit (RFU) for each protein or protein complex for a standard plasma volume per participant was measured with the SomaScan® 5K Assay (SomaLogic). The assay, SomaLogic quality control, and quality control procedures implemented by the ARIC investigators have been described.^21^ Protein RFUs were log_2_transformed and preprocessed using adaptive normalization maximum likelihood (ANML) standardization.^21^ In the main analyses, we used Visit 2 (1990-1992) protein levels. The same procedures were performed for blood collection and protein measurements at Visit 3 (1993-1995) and 5 (2011-2013); data from these visits along with Visit 2 were used for investigating within-person variation in levels over time.

Proteomic profiling produced 4,955 aptamers that passed quality control (out of 5,284), representing 4,712 plasma proteins or protein complexes. We further linearly regressed the aptamer log_2_RFU values on study center, top 10 genetic principal components (PCs), and 10 probabilistic estimation of expression residuals (PEER) factors.^22^ PEER factors adjust for latent patterns associated with technical variation, including uncorrected batch effects. Residuals from this linear regression were then rank-inverse normalized to avoid the influence of extreme values, and were used as the corrected-protein quantification in the analysis.^23^ The preprocessed protein log_2_RFU values for the downstream analyses approximately follow a mean of 0 and a standard deviation (SD) of 1.

Genotyping in ARIC was performed using the Affymetrix Genome-Wide Human SNP Array 6.0, with imputation based on the TOPMed reference panel.^24^ Genetic PCs for ARIC participants were calculated by projecting onto the PC axes derived from a separate, high-quality, multi-ancestry reference dataset, the 1000 Genomes (1000G) + HGDP Project.^25,26^

### Covariate assessment

We selected known and suspected demographic, anthropometric, and lifestyle risk factors for each cancer at Visit 2 (unless otherwise specified) as covariates. Age (years), sex (female or male; Visit 1), race (Black or White; Visit 1), smoking status (current, former, never), packyears, and alcohol drinking status (current, former, never) were assessed by interview. Height and weight, from which body mass index (BMI, kg/m^2^) was calculated, and waist and hip circumferences from which waist-to-hip ratio was calculated, were measured by trained technicians. Physical activity (available only in Visit 1) was assessed using the Baecke questionnaire^27^ for sport, leisure, and work. Diagnosed diabetes (yes, no) was defined by self-report of a physician diagnosis or use of diabetes medication. Among participants without a diabetes diagnosis, pre-diabetes (yes, no) was defined as fasting glucose 100-125 mg/dL (non-fasting 140-199 mg/dL) or HbA1c 5.7-6.4%; undiagnosed diabetes (yes, no) was defined as fasting glucose ≥126 mg/dL (non-fasting ≥200 mg/dL) or HbA1c ≥6.5%; blood levels were measured under ARIC study protocols. We also selected a smoking-related plasma protein score, which we previously developed.^28^

### Statistical analysis

We estimated hazard ratios (HRs) and 95% confidence intervals (CIs) for each cancer site using Cox proportional hazards regression models,^29^ with time since Visit 2 as the underlying time scale. Because our goal was to identify plasma proteins associated with cancer risk beyond known cancer risk factors, in Model 1, we adjusted for common risk factors, including baseline age, a combined race-center variable (all participants recruited in Jackson, MS are Black), sex, smoking status, packyears, BMI, and top 10 genetic PCs. For prostate cancer, sex was excluded from the model. In Model 2, we additionally adjusted for site-specific risk factors described in **Table S2**, and for the top 10 PEER factors. Site-specific risk factors were identified through a comprehensive literature review and were confined to variables accessible within the ARIC study. Adjustment for known cancer risk factors minimizes confounding bias from environmental exposures that differ between incident cases and the general population. Additionally, PEER factors can detect latent artifacts such as batch effects. Together, these adjustments yield more precise risk estimates and robust protein-cancer associations for each site. A Benjamin-Hochberg false discovery rate (FDR) correction^30^ was applied for multiple testing (*FDR* < 0.05).

We conducted sensitivity analyses using time-stratified models to assess how protein–cancer associations varied with time to diagnosis and to confirm whether identified proteins in Model 2 are more likely to reflect cancer etiology and long-term risk rather than the presence of undiagnosed disease. For these analyses, we began follow up >5 years after Visit 2 but used Visit 2 proteins (lagged) or began follow up at Visit 2 and truncated (censored) the follow-up time at 10 years. Proteins that remain associated with risk in the lagged analysis are less likely to represent effects of preclinical or undiagnosed cancer. In the context of risk, truncating the follow-up time may reduce measurement error related to the use of proteins measured once in the setting of within-person variability in protein levels over the long term. To assess the size of any within-person variability, we examined temporal correlations of proteins identified in both models (Models 1 and 2) across multiple study visits. Pearson correlations, and two-way mixed single score intraclass correlations ICC(3,1)^31^ were calculated respectively for ANML standardized log2RFU protein levels between the baseline Visit 2 (1990-1992), Visit 3 (1993-1995), and Visit 5 (2011-2013) in participants with no prevalent cancer at baseline, no incident cancer during follow-up, and complete proteomic data at all 3 visits [N=3,014; N(Male)=1,198; N(Female)=1,816)].

All analyses were conducted using R version 4.2.1 and all tests were conducted as two-sided.

### Pathway enrichment analysis for proteins related to primary liver cancer risk

For primary liver cancer, we conducted enrichment analyses to understand their biological functions. Over-represented Gene Ontology (GO) biological processes, cellular components, and molecular functions were identified using the clusterProfiler R package (v4.4.4).^32^ Reactome pathway enrichment was performed using the ReactomePA package (v1.40.0).^33^ Pathway enrichment analyses were conducted using one-sided hypergeometric testing, with multiple testing correction applied via the Benjamini-Hochberg FDR method. We did not conduct pathway enrichment analyses for the other 6 cancers because the numbers of proteins identified in the full models (Model 2) were insufficient for meaningful analysis.

### Confirmation cohort: the European Prospective Investigation into Cancer and Nutrition (EPIC)

We sought to confirm the proteins associated with the site-specific cancers after FDR correction identified in ARIC in an independent prospective study, the EPIC cohort. The proteomic study in EPIC uses the case-cohort design and includes participants from 4 EPIC countries (United Kingdom, Netherlands, Spain, Italy).^34,35^ Cancer cases were identified over 20 years (10 years median follow-up for cancer cases). Plasma proteins were measured using the SomaScan® 7K Assay v4.1 in blood collected before the cancer diagnoses. All proteins in the ARIC study (5K Assay) are available in the EPIC study (7K Assay). Associations in EPIC were assessed using Prentice-weighted Cox regression^36^ with age as the time scale, adjusting for both common and site-specific risk factors (Model 2) as in ARIC. Note that genetic PCs and PEER factors were not adjusted in the EPIC models due to the cohort consisting of only white participants and different protein measurement and processing procedures.

## Results

### Cohort description

The analytic cohort included 9,391 ARIC participants followed for a median of 22.6 years (range 0.1-25.9 years). Baseline characteristics are shown in **Table 1 and Supplementary Table S1**. A total of 1,617 incident cases of these 7 cancers were diagnosed over a median of 12.1 years (range 0.1-25.2 years). Mean age at Visit 2 was 57 years (SD 5.7). The study population was 54.8% female and 23.3% Black.

**Table 1.**
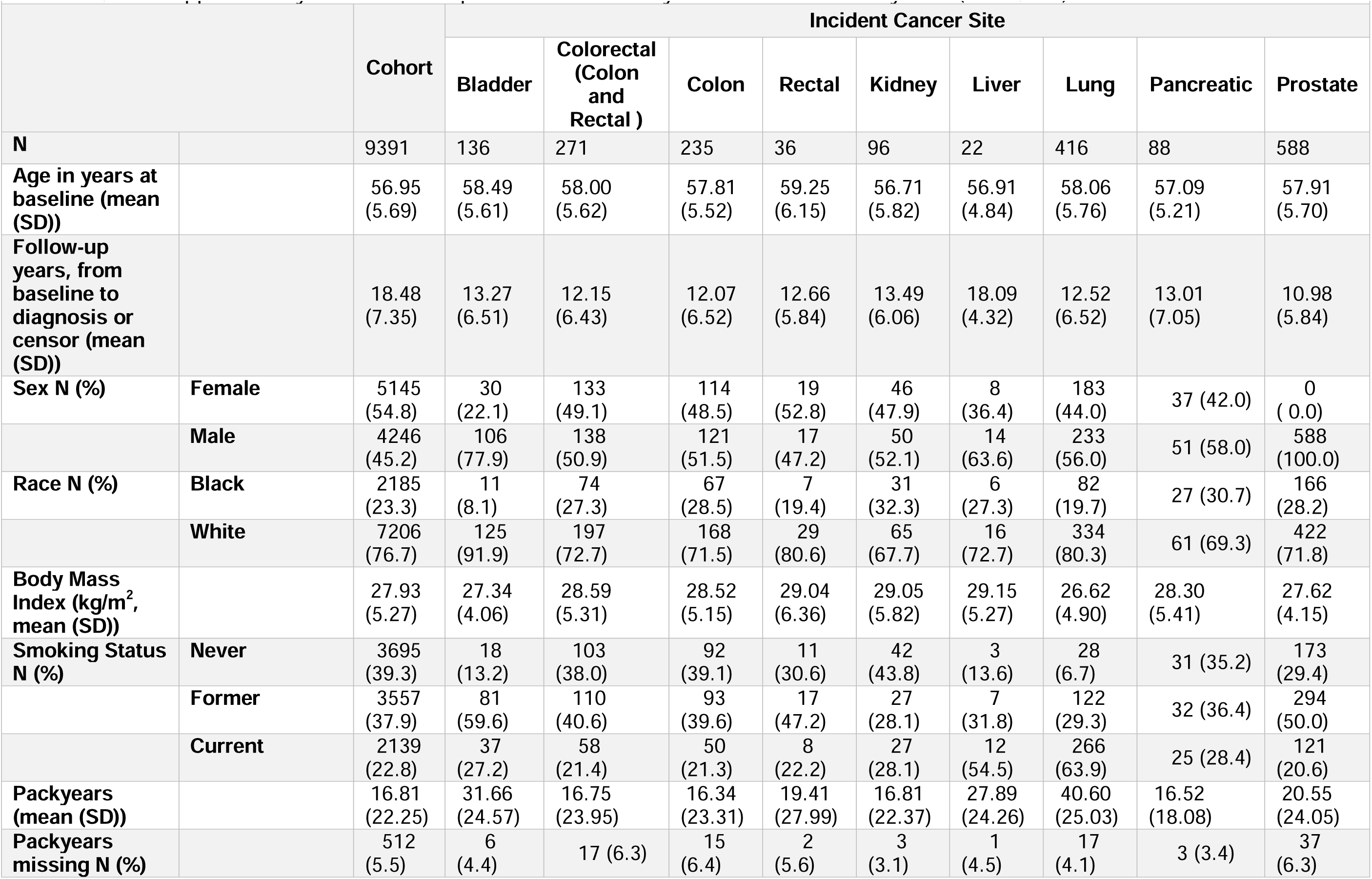
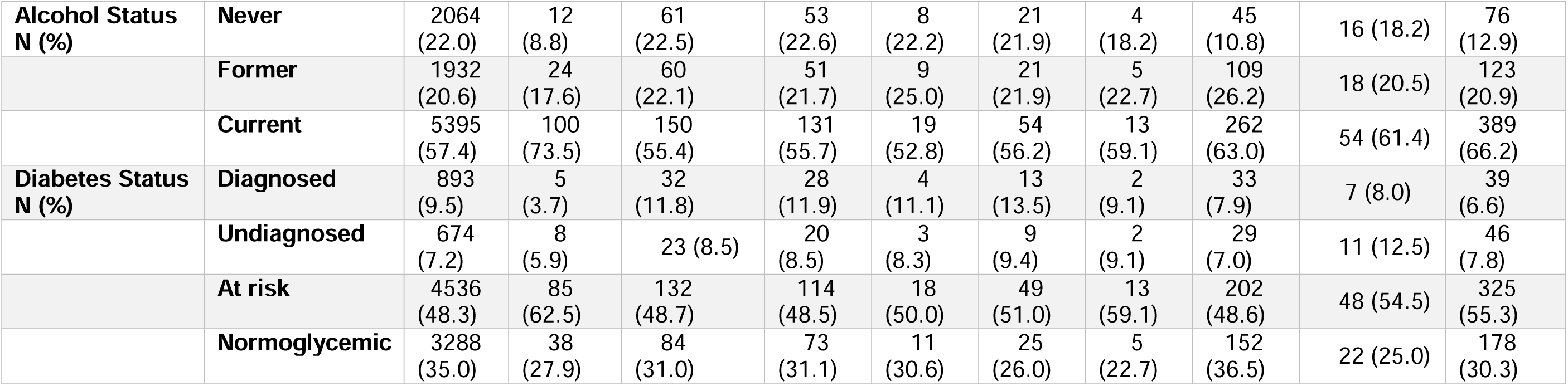
Descriptive characteristics of the analytic cohort and by incident cancer site in ARIC. For the full table and stratified by sex table, see Supplementary Table S1. The prostate cancer analytic cohort included only men (N = 4,246).

|  |  | Cohort | Incident Cancer Site |  |  |  |  |  |  |  |  |
| --- | --- | --- | --- | --- | --- | --- | --- | --- | --- | --- | --- |
|  |  |  | Bladder | Colorectal<br>(Colon<br>and<br>Rectal ) | Colon | Rectal | Kidney | Liver | Lung | Pancreatic | Prostate |
| **N** |  | 9391 | 136 | 271 | 235 | 36 | 96 | 22 | 416 | 88 | 588 |
| **Age in years at baseline (mean (SD))** |  | 56.95<br>(5.69) | 58.49<br>(5.61) | 58.00<br>(5.62) | 57.81<br>(5.52) | 59.25<br>(6.15) | 56.71<br>(5.82) | 56.91<br>(4.84) | 58.06<br>(5.76) | 57.09<br>(5.21) | 57.91<br>(5.70) |
| **Follow-up years, from baseline to diagnosis or censor (mean (SD))** |  | 18.48<br>(7.35) | 13.27<br>(6.51) | 12.15<br>(6.43) | 12.07<br>(6.52) | 12.66<br>(5.84) | 13.49<br>(6.06) | 18.09<br>(4.32) | 12.52<br>(6.52) | 13.01<br>(7.05) | 10.98<br>(5.84) |
| **Sex N (%)** | **Female** | 5145<br>(54.8) | 30<br>(22.1) | 133<br>(49.1) | 114<br>(48.5) | 19<br>(52.8) | 46<br>(47.9) | 8<br>(36.4) | 183<br>(44.0) | 37 (42.0) | 0<br>( 0.0) |
|  | **Male** | 4246<br>(45.2) | 106<br>(77.9) | 138<br>(50.9) | 121<br>(51.5) | 17<br>(47.2) | 50<br>(52.1) | 14<br>(63.6) | 233<br>(56.0) | 51 (58.0) | 588<br>(100.0) |
| **Race N (%)** | **Black** | 2185<br>(23.3) | 11<br>(8.1) | 74<br>(27.3) | 67<br>(28.5) | 7<br>(19.4) | 31<br>(32.3) | 6<br>(27.3) | 82<br>(19.7) | 27 (30.7) | 166<br>(28.2) |
|  | **White** | 7206<br>(76.7) | 125<br>(91.9) | 197<br>(72.7) | 168<br>(71.5) | 29<br>(80.6) | 65<br>(67.7) | 16<br>(72.7) | 334<br>(80.3) | 61 (69.3) | 422<br>(71.8) |
| **Body Mass Index (kg/m<sup>2</sup>, mean (SD))** |  | 27.93<br>(5.27) | 27.34<br>(4.06) | 28.59<br>(5.31) | 28.52<br>(5.15) | 29.04<br>(6.36) | 29.05<br>(5.82) | 29.15<br>(5.27) | 26.62<br>(4.90) | 28.30<br>(5.41) | 27.62<br>(4.15) |
| **Smoking Status N (%)** | **Never** | 3695<br>(39.3) | 18<br>(13.2) | 103<br>(38.0) | 92<br>(39.1) | 11<br>(30.6) | 42<br>(43.8) | 3<br>(13.6) | 28<br>(6.7) | 31 (35.2) | 173<br>(29.4) |
|  | **Former** | 3557<br>(37.9) | 81<br>(59.6) | 110<br>(40.6) | 93<br>(39.6) | 17<br>(47.2) | 27<br>(28.1) | 7<br>(31.8) | 122<br>(29.3) | 32 (36.4) | 294<br>(50.0) |
|  | **Current** | 2139<br>(22.8) | 37<br>(27.2) | 58<br>(21.4) | 50<br>(21.3) | 8<br>(22.2) | 27<br>(28.1) | 12<br>(54.5) | 266<br>(63.9) | 25 (28.4) | 121<br>(20.6) |
| **Packyears (mean (SD))** |  | 16.81<br>(22.25) | 31.66<br>(24.57) | 16.75<br>(23.95) | 16.34<br>(23.31) | 19.41<br>(27.99) | 16.81<br>(22.37) | 27.89<br>(24.26) | 40.60<br>(25.03) | 16.52<br>(18.08) | 20.55<br>(24.05) |
| **Packyears missing N (%)** |  | 512<br>(5.5) | 6<br>(4.4) | 17 (6.3) | 15<br>(6.4) | 2<br>(5.6) | 3<br>(3.1) | 1<br>(4.5) | 17<br>(4.1) | 3 (3.4) | 37<br>(6.3) |
| **Alcohol Status**<br>**N (%)** | **Never** | 2064<br>(22.0) | 12<br>(8.8) | 61<br>(22.5) | 53<br>(22.6) | 8<br>(22.2) | 21<br>(21.9) | 4<br>(18.2) | 45<br>(10.8) | 16 (18.2) | 76<br>(12.9) |
|  | **Former** | 1932<br>(20.6) | 24<br>(17.6) | 60<br>(22.1) | 51<br>(21.7) | 9<br>(25.0) | 21<br>(21.9) | 5<br>(22.7) | 109<br>(26.2) | 18 (20.5) | 123<br>(20.9) |
|  | **Current** | 5395<br>(57.4) | 100<br>(73.5) | 150<br>(55.4) | 131<br>(55.7) | 19<br>(52.8) | 54<br>(56.2) | 13<br>(59.1) | 262<br>(63.0) | 54 (61.4) | 389<br>(66.2) |
| **Diabetes Status**<br>**N (%)** | **Diagnosed** | 893<br>(9.5) | 5<br>(3.7) | 32<br>(11.8) | 28<br>(11.9) | 4<br>(11.1) | 13<br>(13.5) | 2<br>(9.1) | 33<br>(7.9) | 7 (8.0) | 39<br>(6.6) |
|  | **Undiagnosed** | 674<br>(7.2) | 8<br>(5.9) | 23 (8.5) | 20<br>(8.5) | 3<br>(8.3) | 9<br>(9.4) | 2<br>(9.1) | 29<br>(7.0) | 11 (12.5) | 46<br>(7.8) |
|  | **At risk** | 4536<br>(48.3) | 85<br>(62.5) | 132<br>(48.7) | 114<br>(48.5) | 18<br>(50.0) | 49<br>(51.0) | 13<br>(59.1) | 202<br>(48.6) | 48 (54.5) | 325<br>(55.3) |
|  | **Normoglycemic** | 3288<br>(35.0) | 38<br>(27.9) | 84<br>(31.0) | 73<br>(31.1) | 11<br>(30.6) | 25<br>(26.0) | 5<br>(22.7) | 152<br>(36.5) | 22 (25.0) | 178<br>(30.3) |

Compared with the entire analytic cohort, participants who subsequently developed bladder, liver, lung, or prostate cancer were more frequently former or current smokers and exhibited greater cumulative tobacco exposure, as measured in packyears (unadjusted comparisons).

### Proteomic Associations with Solid Cancers

Of the 4,955 aptamers evaluated, 40 unique proteins (40 aptamers) were significantly associated with risk of at least one cancer site after within-site multiple testing correction (FDR < 0.05), yielding a total of 41 protein–cancer associations in Model 2 (adjusted for common and site-specific risk factors) (**Fig.2a, Table 2**). In Model 1 (adjusted for common risk factors only), 139 unique proteins (140 aptamers) were significantly associated with the risk of at least one cancer site after correction for multiple testing, with a total of 144 protein–cancer associations (**Fig.2b, Supplementary Table S3**).

**Table 2.**
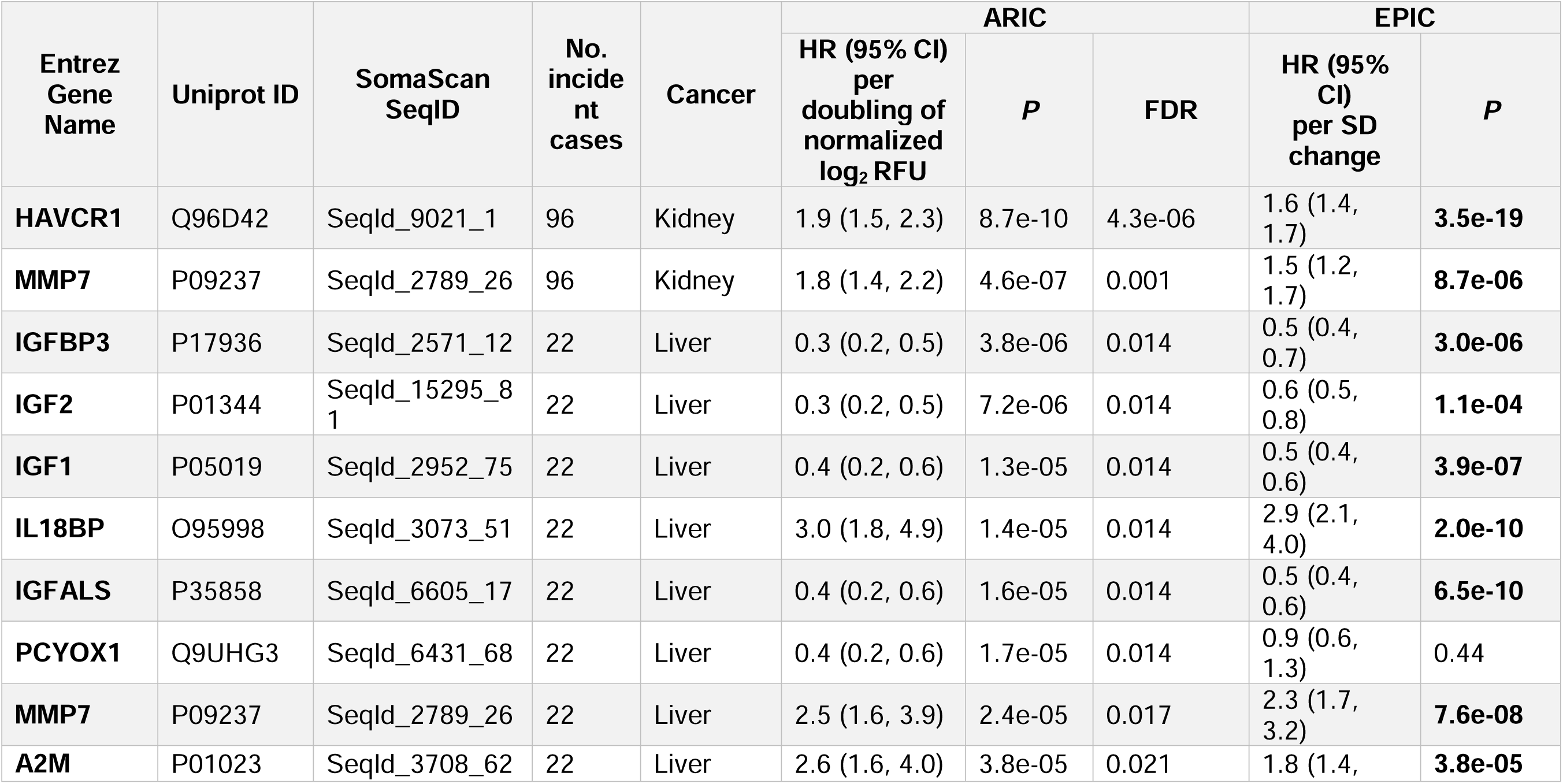
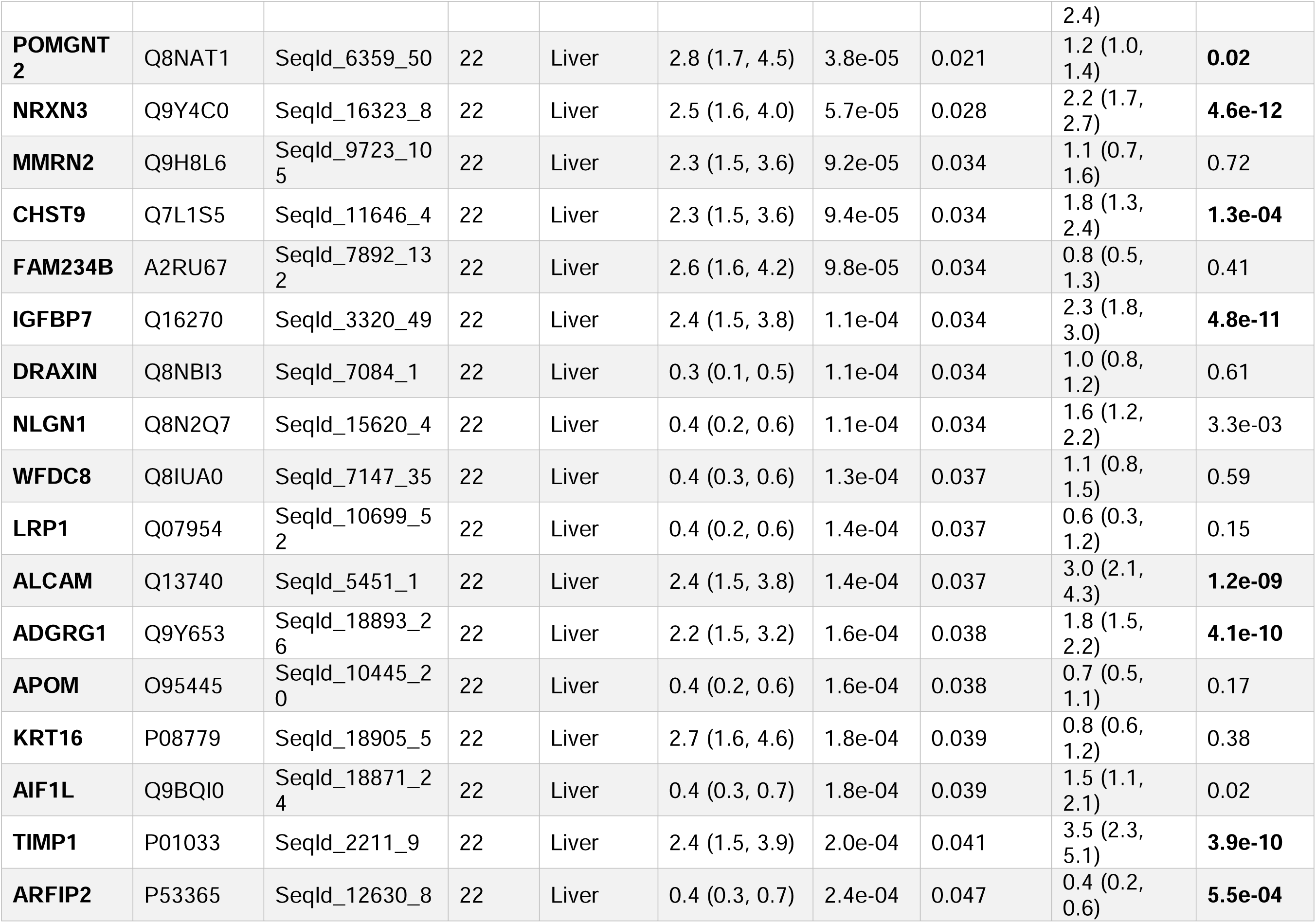
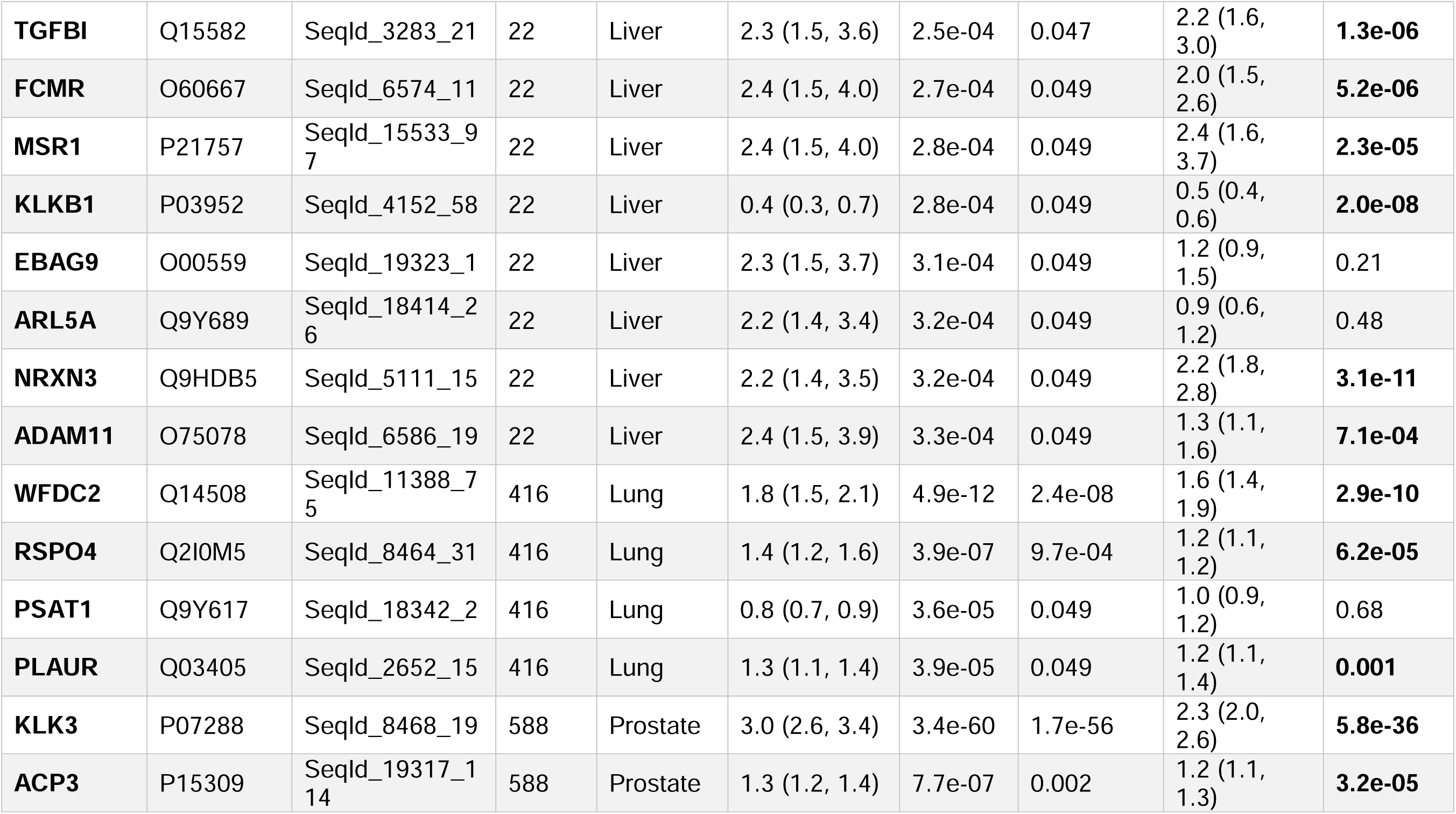
Significant Protein–Cancer Associations Identified in the ARIC Cohort with the EPIC Cohort validations. Hazard ratios (HRs) and 95% confidence intervals (CIs) are from Cox proportional hazards regression analyses adjusted for common and site-specific cancer risk factors (Model 2). For ARIC, both nominal and FDR-adjusted *P* values from two-sided tests are shown. Proteins that met FDR thresholds (0.05) were then evaluated in EPIC. For EPIC, nominal *P* values from two-sided tests are shown. *P* values are displayed in bold when the HRs show consistent directions across the two cohorts and reach nominal significance in EPIC (*P* < 0.05). For all protein associations (Model 2) and the results of models adjusted for common cancer risk factors (Model 1), see Supplementary Tables S6 and S7. Further details of the EPIC cohort validation and comparisons with the published associations in the UK Biobank, which used the Olink panel, are in Supplementary Table S8. Note that in the ARIC results, we present the HR per doubling of normalized log_2_RFU. The preprocessed values are approximately mean 0, SD 1. A strict log(HR) per SD change can be calculated using the SD of protein values provided in Supplementary Table S15 as log(HR) per SD change = log(HR)*SD(protein).

For liver cancer risk, we identified 33 unique protein associations in Model 2, including IGFBP3, IGF2, IGF1, IL18BP, and MMP7 with HRs per doubling (95% CI): 0.3 (0.2, 0.5), 0.3 (0.2, 0.5), 0.4 (0.2, 0.6), 3.0 (1.8, 4.9), and 2.5 (1.6, 3.9), respectively (**Table 2**, **Fig. 3a**). Given that IGFBP3 functions as a carrier protein for IGF1 in circulation, we constructed a joint model including both proteins along with all risk factors. IGFBP3, but not IGF1 remained associated with liver cancer risk [HR (95% CI): IGFBP3, 0.4 (0.2, 1.0); IGF1, 0.8 (0.3, 1.8)]. Two IGF1-targeting aptamers are present in SomaScan (SomaScan Sequence ID: 2952-75 and 8406-17). Only 1 aptamer (SomaScan Sequence ID: 2952-75) demonstrated cancer associations. These 2 IGF1 aptamers were minimally correlated (r = -0.06) in cancer-free participants at Visit 2.

**Figure 3.**
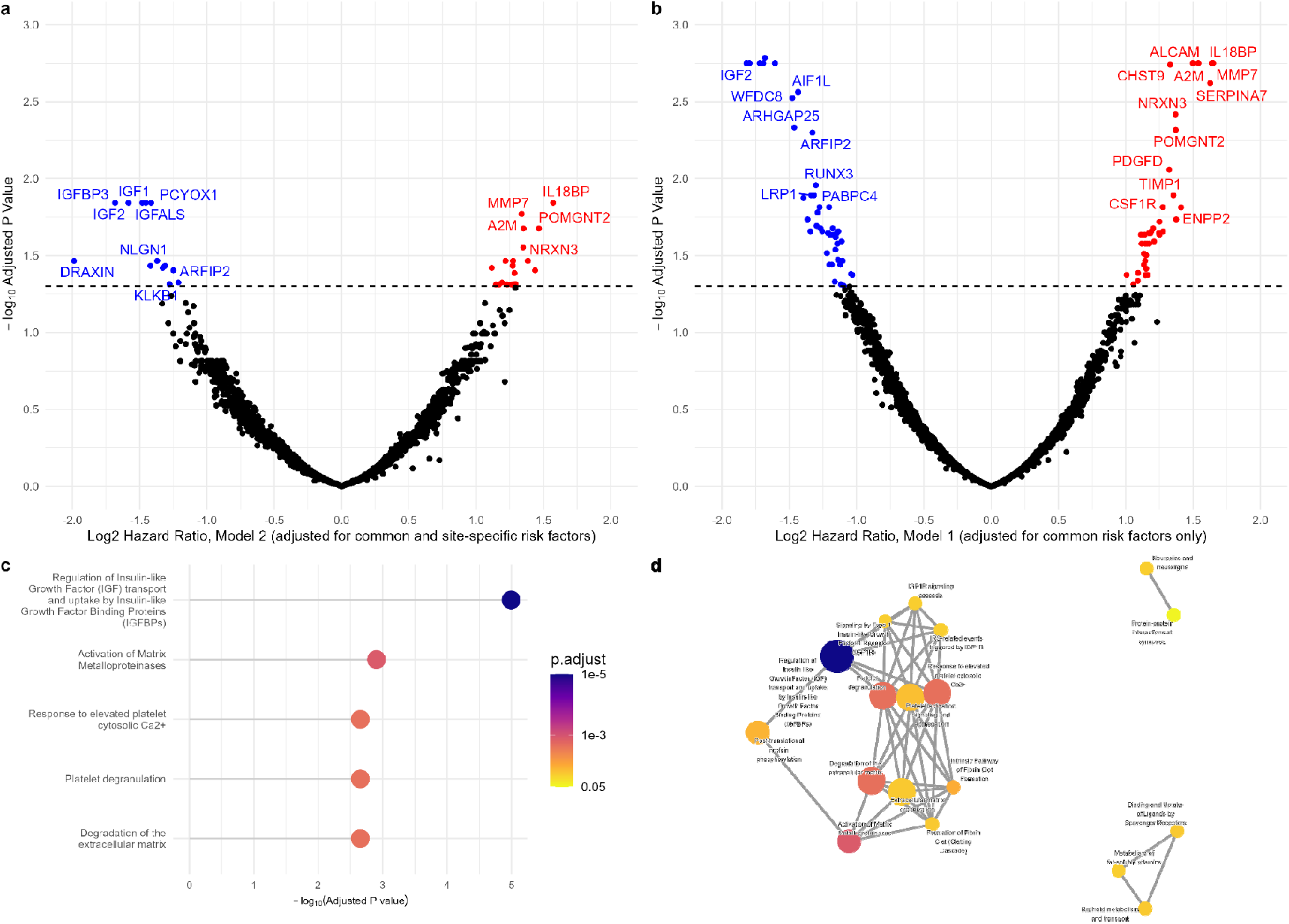
Summary of protein-trait association analysis results for liver cancer in ARIC and biological functions of liver cancer-associated proteins. (a) Volcano plot showing associations between 4,955 aptamers and liver cancer risk from Model 2, adjusted for common and liver cancer-specific risk factors. Log_2_ *hazard ratios* per doubling are plotted on the *x*-axis, –log_10_ FDR-adjusted *P* values are plotted on the *y*-axis. Proteins with FDR < 0.05 are labeled. Hazard ratios and 95% CIs were estimated using two-sided Cox proportional hazards models. Full results in Supplementary Table S7. (b) Volcano plot showing associations between 4,955 aptamers and liver cancer risk from Model 1, adjusted for common risk factors only. Full results in Supplementary Table S6. (c) The top 5 results of Reactome pathway enrichment analysis of 33 proteins related to liver cancer incidence. (d) Enrichment map of Reactome pathways that met the threshold of FDR < 0.05. Connected pathways with edges indicate overlapping gene sets.

Similar results were observed in the joint model of IGFBP3 and IGF2. While both effect sizes were attenuated [HR (95% CI): IGFBP3, 0.5 (0.2, 1.0); IGF2, 0.6 (0.3, 1.3)], IGFBP3 remained more robustly associated with liver cancer risk. To evaluate the influence of liver function on these findings, we performed a sensitivity analysis excluding individuals with potentially impaired hepatic function, defined as those with AST (SomaScan Sequence ID: 4912-17) or ALT (SomaScan Sequence ID: 16015-19) concentrations in the extreme distribution tails (>95th or <5th percentile) at Visit 2. All 33 proteins retained consistent effect directions and statistical significance (**Supplementary Table S4**). In Model 1, 86 aptamers were significantly associated with liver cancer (overlap of 30 aptamers from Model 2). The substantially larger number of proteins identified in Model 1 than in Model 2 suggests that many of the proteins identified in Model 1 are associated with underlying risk factors, such as diabetes and alcohol intake for liver cancer.

For lung cancer risk, 4 proteins were identified: WFDC2, RSPO4, PSAT1, and PLAUR [HR (95% CI): 1.8 (1.5, 2.1)), 1.4 (1.2, 1.6), 0.8 (0.7, 0.9), and 1.3 (1.1, 1.4)]; this model adjusted for smoking status, packyears, and a smoking-related plasma protein score, which reduces the detection of proteins that are smoking-associated by design. Model 1 identified 51 aptamers associated with lung cancer (including all 4 aptamers from Model 2). The larger number of associated proteins may indicate a residual smoking effect for lung cancer.

For prostate cancer risk, 2 proteins were identified in Model 2: KLK3 (prostate-specific antigen) and ACP3 (prostatic acid phosphatase) [HR (95% CI): 3.0 (2.6, 3.4) and 1.3 (1.2, 1.4)]. Model 1 identified the same proteins with similar effect sizes. For kidney cancer, 2 proteins were identified in Model 2: HAVCR1 and MMP7 [HR (95% CI): 1.9 (1.5, 2.3) and 1.8 (1.4, 2.2)]. Sensitivity analysis excluding individuals with poor kidney function at baseline [estimated glomerular filtration rate (eGFR) < 90 and < 60 mL/min/1.73 m², CkdEpi Creatinine-Cystatin Equation 2021], showed consistent results for these 2 proteins (**Supplementary Table S5**). Model 1 also identified the same proteins with similar effect sizes. For colorectal cancer risk, while no proteins met FDR thresholds in Model 2, Model 1 identified 2 proteins associated with colorectal cancer: CLEC3B and BCHE [HR (95% CI): 0.8 (0.7, 0.9) and 0.8 (0.7, 0.9)]. CLEC3B also met the FDR thresholds for colon cancer alone [HR (95% CI): 0.7 (0.6, 0.8)] in Model 1. For bladder and pancreatic cancer risks, no proteins met the FDR thresholds in either Model 1 or Model 2.

Complete results of aptamer-cancer associations for Models 1 and 2 are in **Supplementary Tables S6 and S7**, respectively.

### Confirmation Analyses in the EPIC Cohort

We evaluated FDR-significant protein–cancer associations identified in ARIC in Model 2 (adjusting for both common and site-specific risk factors) in the EPIC study (**Fig. 4**, **Table 2, Supplementary Table S8**). The EPIC study, which used a case–cohort design and focused on White participants only, included a larger number of cancer cases but had shorter follow-up (median: 16 years; median for cases: 10 years). Associations in EPIC were assessed using Prentice-weighted Cox regression^36^ with age as the time scale. In contrast, the ARIC cohort had a substantially longer follow-up (mean: 18.5 years, median: 22.6 years). Proteomic measurements in EPIC (6,412 proteins) and ARIC (4,712 proteins) were both generated using the SomaScan platform.

**Figure 4.**
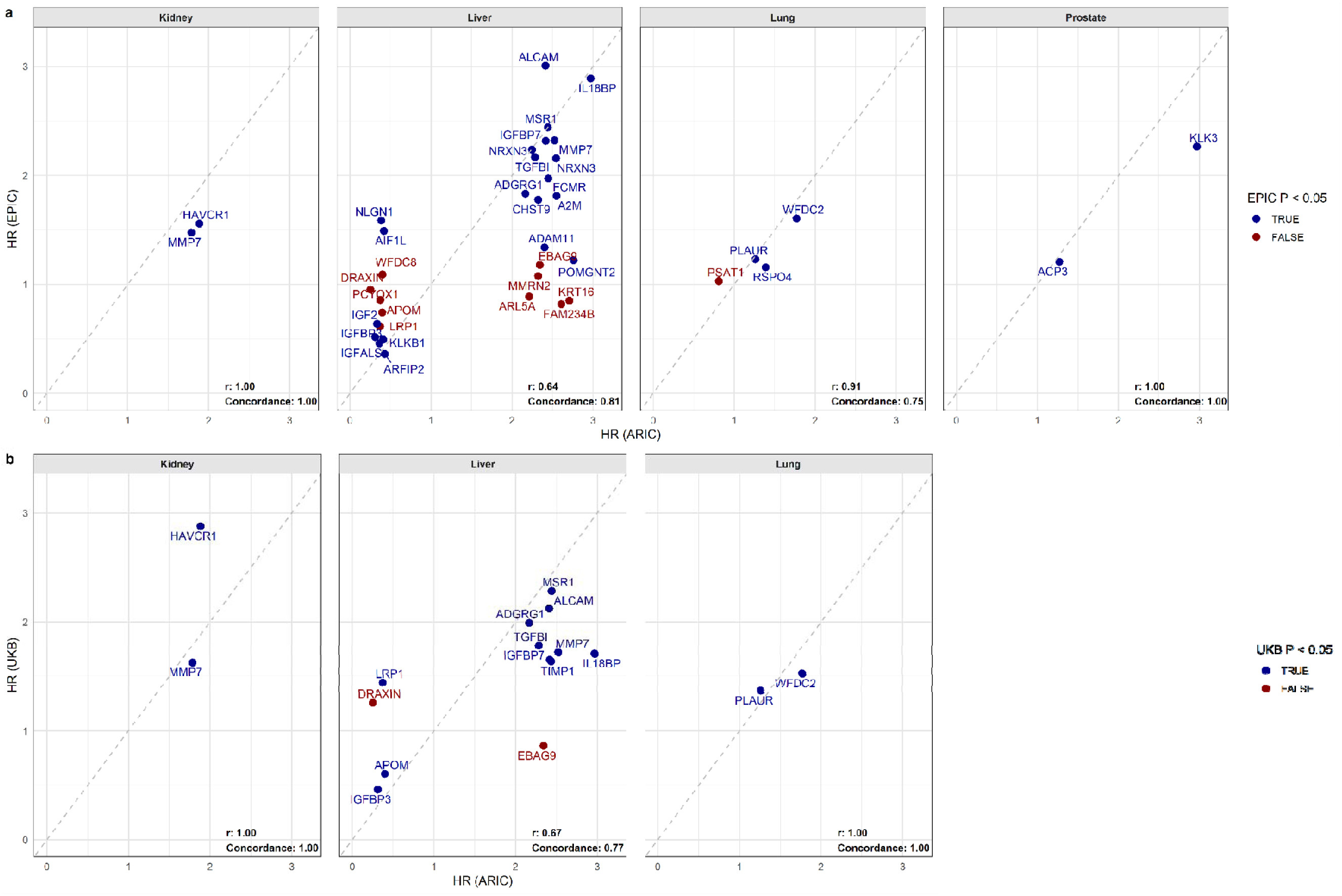
Cross-cohort comparison of FDR-significant proteins in ARIC with confirmation analyses in EPIC (SomaScan) and published results from the UK Biobank (Olink, Papier et al., Nat Commun 2024). a) Hazard ratios (HRs) per doubling for FDR-significant proteins from Model 2 (adjusted for common and site-specific risk factors) in ARIC plotted against HRs in EPIC per SD. All FDR-significant proteins were available in EPIC, with analyses adjusted for the same risk factors as in ARIC. b) HRs per doubling for FDR-significant proteins in ARIC plotted against published HRs per SD from the UK Biobank. UK Biobank used multivariate models adjusted for risk factors, though not entirely identical to ARIC. Of the 40 FDR-significant proteins in ARIC, 16 were available in the UKB, which used the Olink platform. The 2 prostate cancer-associated proteins were not measured in the UK Biobank. Each point represents one protein. Blue dots are FDR significant in ARIC and nominally significant in EPIC or UKB. Red dots are FDR significant in ARIC. Nominal P-values for EPIC and UKB are shown.

Concordance was strong between the FDR-significant protein–cancer associations identified in ARIC and associations for the same proteins estimated in EPIC across all 7 cancer sites adjusting for comparable common and site-specific risk factors as in Model 2 (including the smoking-related plasma protein score for lung cancer) (**Fig. 4**). For liver cancer, 21 of the 33 ARIC-identified proteins were confirmed in EPIC with consistent directions of effects and nominal statistical significance. For lung cancer, 3 of the 4 proteins identified in ARIC (WFDC2, RSPO4, and PLAUR) were confirmed in EPIC, while the 4^th^ protein PSAT1 was not associated with risk [HR (95% CI): 1.0 (0.9, 1.2)]. For prostate cancer, KLK3 and ACP3 were both significant in EPIC with the same direction of association as in ARIC. For kidney cancer, both HAVCR1 and MMP7 demonstrated highly significant associations in EPIC, and both proteins showed consistent association directions across both studies.

### Time-Stratified Analyses

We conducted 2 types of time-stratified analyses using Model 2, which adjusted for both common and site-specific risk factors: a 5-year lagged analysis, in which we began follow up at >5 years after blood draw at Visit 2 (i.e., excluded cases diagnosed or individuals censored ≤5 years after blood draw), and a 10-year truncated analysis, in which we administratively censored follow-up at 10 years after blood draw. Most associations observed in the 5-year lagged analysis were consistent with Model 2 across all 7 cancer sites (**Fig. 5**); 37 of the 41 protein-cancer associations remained FDR significant, and the HRs remained similar to those observed in the analysis starting follow-up at Visit 2. For the prostate cancer protein ACP3, also known as prostatic acid phosphatase (PAP, a known blood-based metastatic prostate cancer marker), the HR was attenuated and was only borderline significant in the lagged analysis, indicating this protein is likely to be an indicator of the presence of an undiagnosed cancer rather than long-term risk. In the 5-year lagged analysis, an additional protein, MGA [HR (95% CI): 0.7 (0.6, 0.8)], met the FDR thresholds for colorectal cancer risk—specifically, colon cancer—despite not being FDR-significant in the full follow-up analysis for Model 2. For liver cancer, all cases occurred after 5 years from baseline at Visit 2, so the results remained the same from the full follow-up model.

**Figure 5.**
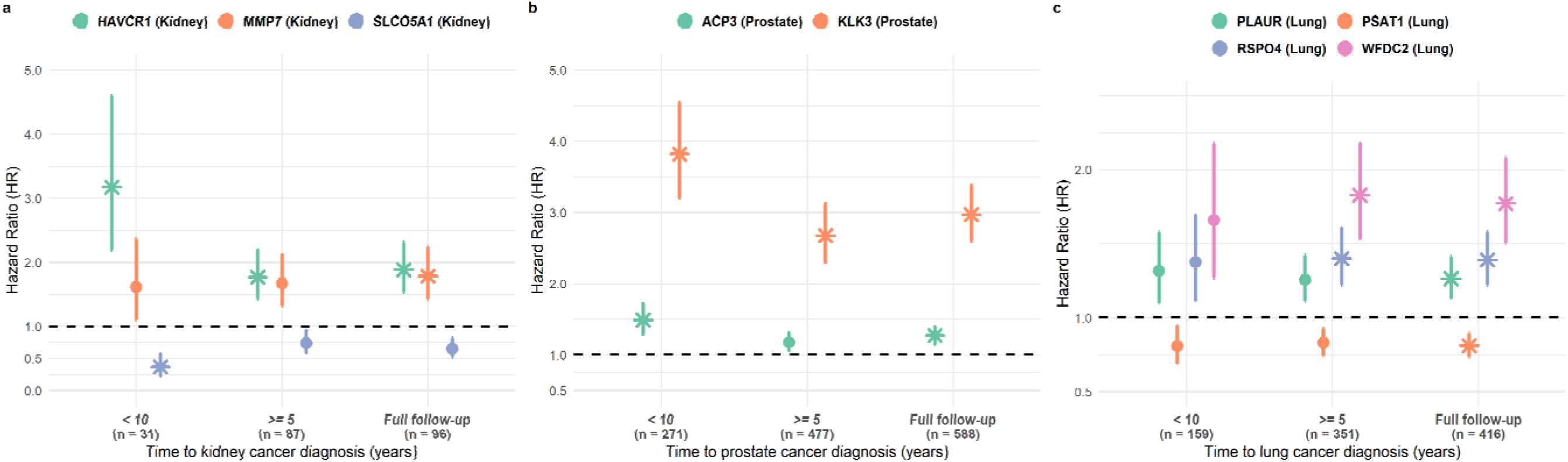
Time-stratified analyses of the association between significant proteins from Model 2 of full follow-up and risk of kidney, prostate, and lung cancer in ARIC. Asterisks (*) denote proteins that remained significant after correction for multiple testing (FDR < 0.05) in the specific time-course analysis. Hazard ratios (HR) per doubling are shown for each protein when follow-up was censored at 10 years (< 10 years) since Visit 2 or lagged by 5 years (>5 years) since Visit 2. HRs from the full follow-up are shown for comparison. All HRs are from Cox models adjusted for common and site-specific risk factors (Model 2). n indicates the number of cases in each analysis. SLCO5A1 for kidney cancer is included in this figure as it emerged as FDR-significant in the truncated 10-year analysis. Complete results for all aptamers in time-stratified analyses are provided in Supplementary Tables S9 and S10. Liver cancer is not shown because all incident cases were diagnosed after 5 years and only 1 case was diagnosed within 10 years since Visit 2.

In the 10-year truncated analysis, we observed changes in the magnitude of the HR for several proteins, including ACP3 and KLK3 for prostate cancer, and HAVCR1 for kidney cancer, for which the associations were stronger compared to the overall analysis. An additional protein, SLCO5A1 [HR (95% CI): 0.4 (0.2, 0.6)], met the FDR thresholds in the truncated 10-year analysis for kidney cancer. For lung cancer, however, the HRs for all the identified proteins remained similar in the truncated analysis compared to the overall analysis, indicating these proteins contain information on risk beyond 10 years. Complete results for all cancers for time-stratified analyses are in **Supplementary Tables S9 and S10**.

### Temporal Variation of Protein Levels

To inform the time-stratified analyses that used proteins that measured once in time, we assessed the long-term variability in protein levels by estimating pairwise correlations and intraclass correlations (ICCs) of the proteins identified in both models (Models 1 and 2) across Visit 2 (1990-1992), Visit 3 (1993-1995), and Visit 5 (2011-2013) among individuals free of prevalent and incident cancer throughout the study period until 2015. Among the 40 proteins identified in Model 2 (adjusted for common and site-specific risk factors), the correlation of protein values between Visits 2 and 3 (Median [Q1,Q3]: 0.6 [0.5, 0.7]) is only slightly larger than the correlations between Visits 2 and 5 (Median [Q1,Q3]: 0.5 [0.4, 0.6]) (**Supplementary Fig. 1**), despite Visits 2 and 3 being only 3 years apart, while Visits 2 and 5 are 21 years apart. The ICCs among all 3 visits of these 40 proteins showed a similar result (Median [Q1,Q3]: 0.5 [0.4, 0.6]). Individual protein trajectories across study visits revealed that the majority of proteins exhibited temporal trends, with levels either increasing or decreasing over time (**Supplementary Fig. 2**). The highest temporal stability was observed for PLAUR (r = 0.8 between Visits 2 and 5; ICC = 0.7 among Visits 2, 3, and 5), 60% (24/40) of proteins showed a correlation greater than 0.5 between Visits 2 and 5, and 58% (23/40) of proteins showed an ICC greater than 0.5 among all 3 visits, indicating consistency over the 21-year observation period. Detailed results of the temporal correlations and ICCs of top proteins identified in both Models 1 and 2 across visits are in **Supplementary Table S11**.

### Pathway enrichment analysis for primary liver cancer

Pathway enrichment analysis of the 33 proteins associated with primary liver cancer risk in Model 2 (FDR < 0.05) revealed significant over-representation of pathways (**Fig. 3c**) related to *insulin-like growth factor (IGF) signaling, extracellular matrix remodeling,* and *platelet activation*. The most significantly enriched pathway was *regulation of IGF transport and uptake by IGFBPs* (FDR = 1.01×10^-5^), involving key proteins such as IGFBP3, IGFBP7, IGF1, IGF2, TIMP1, and IGFALS. Other enriched pathways included *activation of matrix metalloproteinases, platelet degranulation, and degradation of the extracellular matrix*, implicating proteins like MMP7, TIMP1, and A2M.

Network analysis of enriched Reactome pathways among the 33 proteins associated with liver cancer risk revealed three major functional modules (**Fig. 3d**). Module 1, which was the largest cluster, included *IGF signaling, extracellular matrix remodeling, and platelet activation*, suggesting the role for these pathways in the etiology of liver cancer. This was supported by GO enrichment in *negative regulation of cell migration, peptidase activity, growth factor binding, and platelet alpha granules*. Module 2 comprised pathways related to lipid and vitamin metabolism, including *binding and uptake of ligands by scavenger receptors, retinoid metabolism and transport, and metabolism of fat-soluble vitamins*, implicating metabolic dysregulation in liver cancer etiology. This was also supported by GO enrichment for lipoprotein transport, cholesterol/sterol transport, and localization to lipoprotein particles. Module 3 included *neurexins and neuroligins and protein-protein interactions at synapses*, aligning with GO terms such as neuron cell-cell adhesion and amyloid-beta binding, indicating potential links between synaptic-like adhesion mechanisms and liver tumorigenesis. Collectively, these results highlight integrated roles of growth signaling, tissue remodeling, hemostasis, and metabolism in liver cancer risk. All significantly enriched Reactome and GO pathways (FDR < 0.05) are reported in **Supplementary Table S12**.

## Discussion

We report a comprehensive proteomic analysis with the extensive follow-up (25 years) in a U.S. cohort of White and Black participants to evaluate associations between 4,712 circulating proteins (4,955 aptamers), measured on the SomaScan® 5K platform, and the risk of 7 site-specific cancers. Using a conservative approach with detailed accounting for known cancer and site-specific risk factors, we identified 41 plasma proteins significantly associated with cancer risk, of which 39 were linked to cancers diagnosed more than 5 years after blood draw. The majority of these associations were supported by findings from an independent cohort using the same proteomic platform (EPIC, SomaScan). Additional sensitivity analyses showed that 4 proteins were strongly associated with cancers diagnosed within 10 years after blood draw: 2 proteins for kidney cancer risk, HAVCR1 and SLCO5A1, are novel plasma markers; and the 2 proteins for prostate cancer risk KLK3 (PSA) and ACP3 (PAP) are known blood-based biomarkers. KLK3, an established marker for the early detection of prostate cancer when measured using immunoassays,^37^ showed a robust association with prostate cancer risk when measured by SomaScan in our analysis. ACP3 is not currently used for early detection but serves as a plasma biomarker for metastatic prostate cancer.^38^

Our approach enabled us to identify circulating protein biomarkers of cancer risk that provide information beyond established risk factors, including demographic, anthropometric measurements (e.g., BMI), and self-reported lifestyle variables (e.g., smoking history). Models adjusted for common cancer risk factors identified substantially more proteins for liver and lung cancers than models further adjusted for site-specific risk factors, suggesting confounding by diabetes and alcohol intake for liver cancer, and residual confounding by smoking behavior for lung cancer. In contrast, for kidney cancer and prostate cancer, the identified proteins were the same across the two models, and the HRs for these proteins remained unchanged between models, indicating robust associations independent of known site-specific risk factors. The proteins that were associated with risk after adjusting for both common and site-specific risk factors may reflect exposures to unknown risk factors, genetic and epigenetic risk factors, or that those proteins may better capture the biologically effective dose of exposure to the known risk factors than other conventional measurements, as we previously documented for smoking versus smoking-related proteins and lung cancer risk.^28^

Despite the small number of first primary liver cancer cases in ARIC, this site had the highest number of protein associations. The liver is a major source of circulating proteins,^39^ and the integrated pathway enrichment analyses of these significant proteins point to three interconnected molecular modules potentially involved in liver cancer development. Module 1 reinforces known cancer hallmarks, integrating IGF signaling, extracellular matrix (ECM) remodeling, and coagulation, with functional annotations highlighting suppressed cell motility, peptidase inhibition, and enrichment in platelet granules. We previously reported that IGFBP7, which has now been identified as being associated with liver cancer risk in the current study, was positively linked to overall cancer mortality, particularly lung cancer mortality, in the ARIC study^40^. Module 2 emphasizes the contribution of altered lipid and vitamin metabolism—supported by both Reactome and GO pathways—involving scavenger receptors and plasma lipoproteins, which may influence tumor-promoting inflammation and metabolic stress. The synapse-related module (Module 3), while traditionally associated with neuronal function, may reflect aberrant cell–cell adhesion mechanisms mediated by neurexins and neuroligins that have emerging roles in non-neuronal tissue homeostasis and cancer.

Although most of the identified protein-cancer associations were cancer site-specific, we identified some proteins that mark common processes across cancer sites and other proteins with associations specific to a particular cancer site. MMP7, an enzyme involved in breaking down extracellular matrix, playing a role in normal tissue remodeling and pathological processes, was associated with an increased risk of cancers of the liver and kidney in both Model 1 (adjusted for common risk factors only) and Model 2 (adjusted for common and cancer site-specific risk factors). Additionally, in Model 1, TREM2 and ALCAM were associated with both liver and lung cancer risks (FDR<0.05). **Supplementary Tables S13 and S14** summarize proteins in common across cancer sites using a more liberal significance threshold, and specify the expected number of false positives.

Protein–cancer risk associations were most common for cancers arising in blood filtration organs, such as the liver and kidney, and these associations remained after excluding participants with extreme levels of liver and kidney function markers. In contrast, fewer associations were identified for cancers of the prostate, bladder, colorectum, and pancreas. This pattern may reflect several factors: these complex diseases may involve many proteins with small effect sizes that are difficult to detect given current sample sizes; key risk-associated exposures or biological processes for these cancer types may not be well-captured by the circulating proteins (both secreted and those that leak into circulation) measured in the current high-throughput proteome panel, and/or that some organs produce proteins locally that are not secreted into the circulation.^41^

In time-stratified analyses, most protein–cancer associations remained robust over time, supporting their relevance for long-term risk prediction. In the 5-year lagged analysis, 37 of 41 FDR-significant associations from the full model remained FDR significant, with one additional protein (MGA) emerging as significant for colon cancer (and combined colorectal cancer). MGA is a transcription factor and member of the MYC regulatory network that is frequently mutated or deleted across multiple cancer types, including colon cancer.^42^ The circulating form of MGA identified in this study may show evidence as a biomarker for colon cancer risk prediction. For prostate cancer, KLK3 was consistently associated across time windows, while ACP3 met FDR thresholds in the 10-year truncated model and not in the 5-year lagged analysis, suggesting it may be a shorter-term marker of risk—consistent with findings from the EPIC cohort. For kidney cancer, HAVCR1 showed stronger associations in the truncated 10-year follow-up period and stable associations in the lagged analysis similar to the full follow-up, indicating potential utility as a marker for both near-term and long-term risk. Temporal correlation analyses demonstrated robust long-term within-person stability of HAVCR1, further supporting its potential as a biomarker for kidney cancer risk prediction.

In addition to the EPIC cohort, we sought to further confirm our findings by evaluating the concordance of the 40 FDR-significant proteins (41 protein-cancer associations) identified in ARIC in a cohort using a different proteomic platform. We compared our results with published findings in the UK Biobank, which employed Olink technology to measure 2,074 proteins in plasma samples from 54,306 middle-aged adults.^15^ Of our ARIC-identified proteins, 16 proteins (17 protein-cancer associations) were available in the UK Biobank (comparison of findings in **Fig. 4** and **Supplementary Table S8**). While the UK Biobank cohort provided a larger sample size, it had a shorter mean follow-up period (12 years) compared to ARIC. For kidney cancer, both HAVCR1 and MMP7 demonstrated highly significant associations in the UK Biobank, with consistent directions. Similarly, for lung cancer, both available proteins (WFDC2 and PLAUR) exhibited significant associations with consistent directions between cohorts. Most notably, for liver cancer, 10 of the 13 ARIC-identified proteins available in the Olink panel were also identified in the UK Biobank study. The reproducibility of these protein-cancer associations across 3 independent cohorts with different demographic characteristics and distinct proteomic platforms (2 SomaLogic vs. 1 Olink) strengthens confidence in our findings. These cross-platform and cross-population confirmations are critical given the known technical differences between proteomic assays^11^ and suggest that these biomarkers may have robust clinical utility for long-term cancer risk stratification.

This study has a number of strengths. Proteomic measurements were obtained at Visit 2 for the majority of ARIC participants, providing a robust baseline for prospective analyses. The extended follow-up period enabled the assessment of circulating proteins as risk factors for cancer incidence, rather than markers of early detection. The availability of protein measurements at 3 timepoints spanning 21 years allowed us to evaluate long-term within-person temporal stability of protein levels to inform future studies. Additionally, the population diversity of the ARIC cohort (25% Black, 75% White participants) enhances the generalizability of these findings to broader populations. The study benefits from high-quality data, including adjudicated cancer cases primarily from cancer registry linkage, comprehensive data on established cancer risk factors, and minimal missing covariate information. Furthermore, we confirmed our findings in an independent cohort that used the same proteomic platform (SomaScan), and in which we harmonized to have comparable covariate adjustments. We also compared our findings with published findings from another independent cohort that used a different technology (Olink).

This study also had some limitations. In ARIC, the number of incident cancer cases was modest for most sites, which limits our ability to identify weaker associations at a stringent significance level accounting for multiple testing. Nevertheless, proteins were statistically significant after p-value adjustment for false discovery, and most were confirmed in 2 independent studies. We did not assess the possibility of non-linear associations. We studied 7 site-specific cancers, and we previously published on breast cancer^17^ and served as a confirmation cohort for lymphomas^18^; other cancer sites will require longer follow-up or pooling with other cohort studies. Finally, protein measurements were obtained when participants were middle-aged; it remains unclear whether results would generalize to measurements obtained at younger or older ages.

In conclusion, we identified multiple circulating plasma proteins associated with cancer risk using data from both Black and White participants in the ARIC study, with confirmation in the EPIC cohort and concordance with published findings in the UK Biobank using a different proteomics platform. In ARIC, many of these associations in the risk factor-adjusted models were observed more than 5 years prior to cancer diagnosis, suggesting that these factors may manifest long before diagnosis and reflect biological phenomena that contribute to the disease’s etiology. These associations were observed after adjusting for common and site-specific cancer risk factors, suggesting that these proteins may reflect a complex mixture of downstream effects of unknown and residual known factors that contribute to cancer development. Future mechanistic investigations are needed to establish causal relationships for etiology using methods such as Mendelian randomization and genetically predicted protein-cancer risk associations.^16,43,44^ Future studies are also needed to evaluate the utility of the identified proteins for cancer risk stratification, with or without measurements on known risk factors. Validated protein biomarkers could ultimately contribute to the development of novel interventions and improved risk prediction for targeted primary and secondary cancer prevention.

## Data Availability

All data produced in the present study are available upon reasonable request to the authors

## Acknowledgments

*ARIC.* The Atherosclerosis Risk in Communities study has been funded in whole or in part with Federal funds from the National Heart, Lung, and Blood Institute, National Institutes of Health, Department of Health, and Human Services, under Contract nos. (75N92022D00001, 75N92022D00002, 75N92022D00003, 75N92022D00004, 75N92022D00005). The authors thank the staff and participants of the ARIC study for their important contributions. Studies on cancer in ARIC are also supported by the National Cancer Institute (U01 CA164975: PI Elizabeth Platz) and the National Heart, Lung, and Blood Institute, National Institutes of Health, Department of Health, and Human Services, under Contract nos. (75N92022D00001, 75N92022D00002, 75N92022D00003, 75N92022D00004, 75N92022D00005). The authors thank the staff and participants of the ARIC study for their important contributions. The content of this work is solely the responsibility of the authors and does not necessarily represent the official views of the National Institutes of Health.

Cancer incidence data have been provided by the Maryland Cancer Registry, Center for Cancer Surveillance and Control, Department of Mental Health, and Hygiene, 201 W. Preston Street, Room 400, Baltimore, MD 21201. We acknowledge the State of Maryland, the Maryland Cigarette Restitution Fund, and the National Program of Cancer Registries (NPCR) of the Centers for Disease Control and Prevention (CDC) for the funds that helped support the availability of the cancer registry data.

SomaLogic Inc. conducted the SomaScan assays in exchange for use of ARIC data. This work was supported in part by NIH/NHLBI grant R01 HL134320. All papers will be shared with SomaLogic by the ARIC publications committee.

Funding was also supported by R01HL087641 and R01HL086694; National Human Genome Research Institute contract U01HG004402; and National Institutes of Health contract HHSN268200625226C. Infrastructure was partly supported by Grant Number UL1RR025005, a component of the National Institutes of Health and NIH Roadmap for Medical Research.

*EPIC*. The authors thank all study subjects for their participation and all interviewers who participated in the fieldwork studies in each EPIC center. The authors also thank Bertrand Hemon at IARC for his valuable work and technical support with the EPIC database. The coordination of EPIC-Europe is financially supported by International Agency for Research on Cancer (IARC) and also by the Department of Epidemiology and Biostatistics, School of Public Health, Imperial College London which has additional infrastructure support provided by the NIHR Imperial Biomedical Research Centre (BRC). The national cohorts are supported by Associazione Italiana per la Ricerca sul Cancro-AIRC-Italy, Italian Ministry of Health, Italian Ministry of University and Research (MUR), Compagnia di San Paolo (Italy); Dutch Ministry of Public Health, Welfare and Sports (VWS), the Netherlands Organisation for Health Research and Development (ZonMW), World Cancer Research Fund (WCRF), (The Netherlands); Instituto de Salud Carlos III (ISCIII), Regional Governments of Andalucía, Asturias, Basque Country, Murcia and Navarra, and the Catalan Institute of Oncology - ICO (Spain); Cancer Research UK (C864/A14136 to EPIC-Norfolk; C8221/A29017 to EPIC-Oxford), Medical Research Council (MR/N003284/1, MC-UU_12015/1 and MC_UU_00006/1 to EPIC-Norfolk; MR/Y013662/1 to EPIC-Oxford) (United Kingdom). Previous support has come from “Europe against Cancer” Programme of the European Commission (DG SANCO). The funders were not involved in designing the study; collecting, analyzing or interpreting the data; or in writing or submitting the manuscript for publication.

*Trans-Omics in Precision Medicine (TOPMed)* program imputation panel (version Freeze5) was supported by the National Heart, Lung and Blood Institute (NHLBI); see www.nhlbiwgs.org. TOPMed study investigators contributed data to the reference panel, which can be accessed through the Michigan Imputation Server; see https://imputationserver.sph.umich.edu. The panel was constructed and implemented by the TOPMed Informatics Research Center at the University of Michigan (3R01HL-117626–02S1; contract HHSN268201800002I). The TOPMed Data Coordinating Center (3R01HL-120393–02S1; contract HHSN268201800001I) provided additional data management, sample identity checks, and overall program coordination and support. We gratefully acknowledge the studies and participants who provided biological samples and data for TOPMed.

## Ethics approval and consent to participate

*ARIC.* Approval for the ARIC study was received from the Institutional Review Board at each study center. All participants provided informed consent.

*EPIC*. The EPIC study was conducted in accordance with the Declaration of Helsinki. The study was approved by the local ethical committees in participating countries and the IARC ethical committee. All participants provided written informed consent for data collection and storage, as well as individual follow-up before study entry. This study is listed at clinicaltrials.gov as NCT03285230.

## IARC Disclaimer

Where authors are identified as personnel of the International Agency for Research on Cancer / World Health Organization, the authors alone are responsible for the views expressed in this article and they do not necessarily represent the decisions, policy or views of the International Agency for Research on Cancer / World Health Organization.

## Data Availability Statement

*ARIC*. Pre-existing data access policies for the parent ARIC cohort study specify that research data requests can be submitted to the steering committee; these will be promptly reviewed for confidentiality or intellectual property restrictions and will not be refused unreasonably. Individual level patient or protein data may further be restricted by consent, confidentiality, or privacy laws/considerations. These policies apply to both clinical and proteomic data. Additional information on how to obtain data, including access to some data via BioLINCC, which is free of charge and without the need for the ARIC Study approval, is available at: https://aric.cscc.unc.edu/aric9/researchers/Obtain_Submit_Data.

*EPIC*. For information on how to submit an application for gaining access to EPIC data and/or biospecimens, please follow the instructions at https://login.research4life.org/tacsgr0epic_iarc_fr/access/index.php.

