## Supplemental Figures for "Identifying Plasma Proteins Associated with Risk of Solid Cancers: A 25-Year Prospective Analysis of 4,712 Circulating Proteins in the ARIC Study"

**Supplementary Figure 1. Temporal correlations of cancer risk-associated proteins across study visits in participants who remained cancer-free.** Correlations between ANML standardized log2(RFU) protein levels measured at Visit 2 (1990-1992), Visit 3 (1993-1995), and Visit 5 (2011-2013) in participants who remained cancer-free throughout follow-up (N=3,014). Cancer risk-associated proteins were identified from Model 2, adjusted for common and site-specific risk factors. Analysis restricted to participants with no prevalent cancer at baseline, no incident cancer during follow-up, and complete proteomic data at all 3 visits. Due to their prostate-specific nature, analyses for KLK3 and ACP3 are conducted among male participants only (N = 1,198).

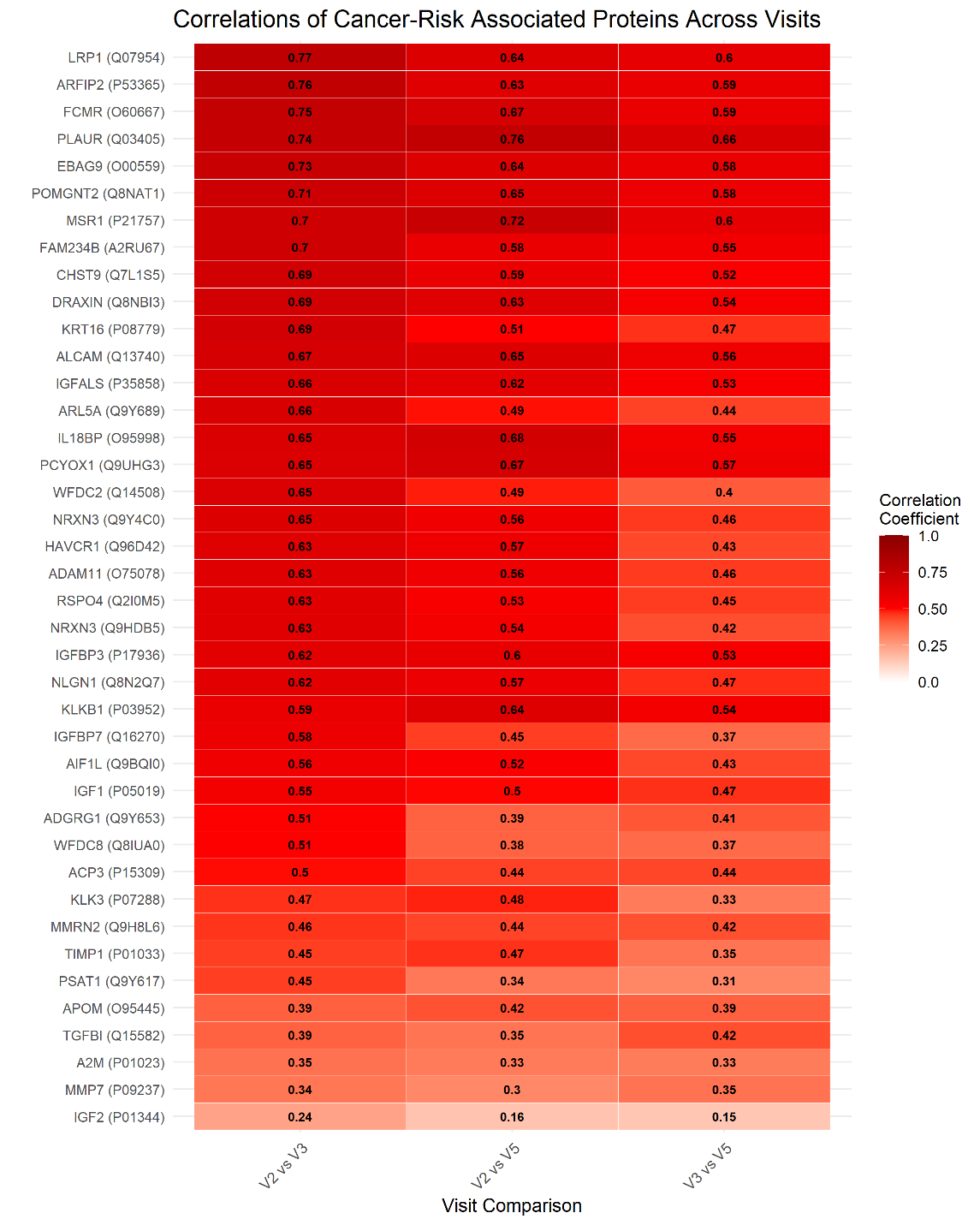

**Supplementary Figure 2. Longitudinal trajectories of cancer risk-associated proteins in participants who remained cancer-free.** Protein concentration profiles over time (Visit 2, Visit 3, and Visit 5) for individuals who remained cancer-free throughout the study period (N = 3,014). Proteins are selected based on the results in Model 2, adjusted for common and site-specific cancer risk factors. Participants had no prevalent cancer at baseline and no incident cancer diagnosis during follow-up, with complete proteomic measurements available at all 3 study visits. Red line with dots represents mean protein values across visits for 3,014 individuals. Dark blue box plots show median, Q1, and Q3 protein values at each visit. Light blue lines represent individual protein trajectories across visits. Due to their prostate-specific nature, protein values for KLK3 and ACP3 are presented among male participants only (N = 1,198).

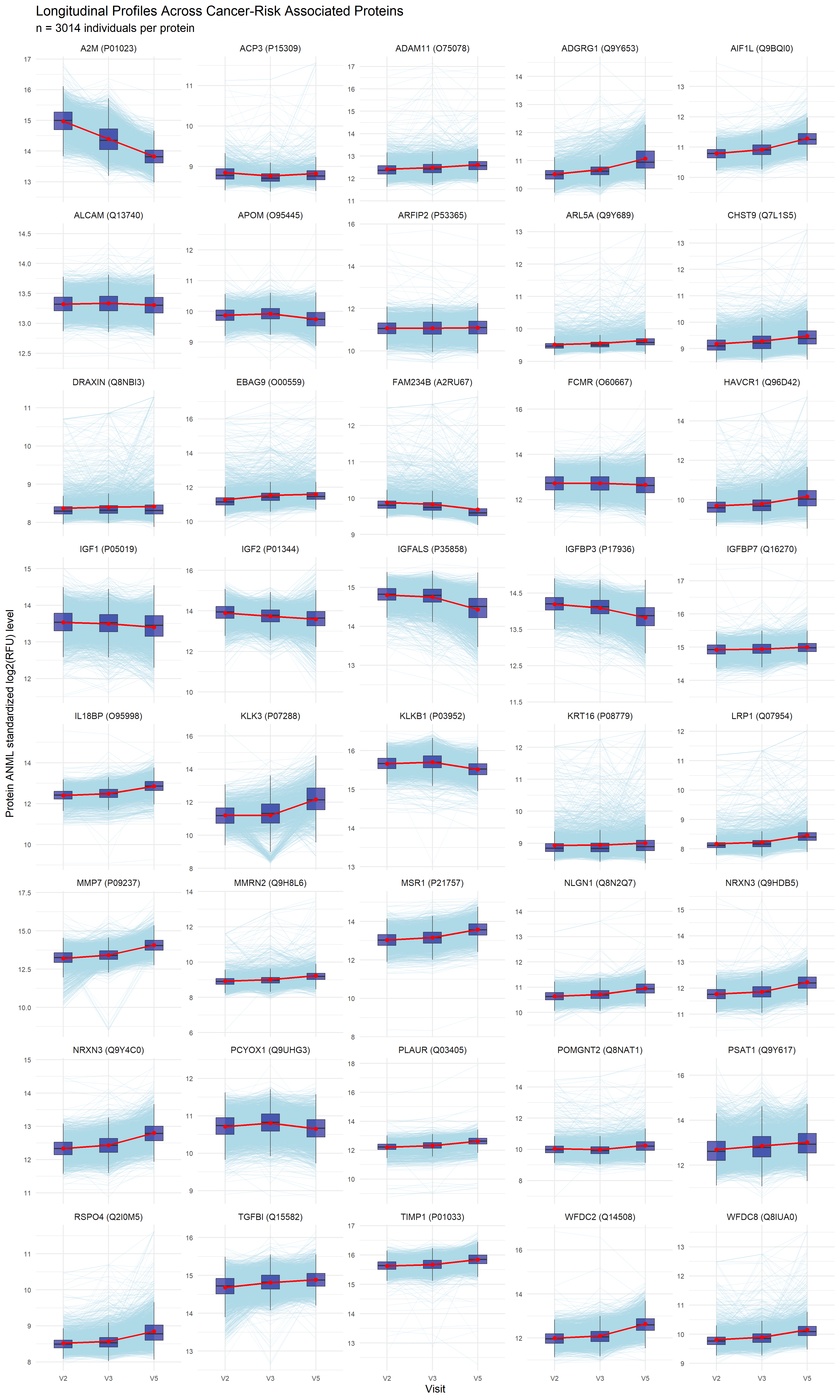
